# Cancer Survival at a Comprehensive Cancer Center Compared with Surveillance, Epidemiology, and End Results (SEER) Estimates

**DOI:** 10.1101/2025.09.05.25335204

**Authors:** Ravi Salgia, Joseph Alvarnas, Vijay Trisal, Ann Vanderplas, Xiaochen Li, Javier Arias-Romero, Jeremy Fricke, Paul Frankel, Isa Mambetsariev, Hannah Rice, Harlan Levine, Marcel R.M. van den Brink

## Abstract

**Background:** Cancer Centers offer a comprehensive approach to cancer prevention, diagnosis, and treatment through tertiary services and by providing coordinated, personalized care. We investigated whether this approach results in significantly better patient outcomes at an NCI-designated Comprehensive Cancer Center (NCI-CCC), City of Hope (COH), versus the U.S. Surveillance, Epidemiology, and End Results (SEER) national database.

**Methods:** Patient-level data were abstracted from the COH Cancer Registry and SEER*Stat software, respectively. The cohort included patients diagnosed with incident cancer from 2004 to 2020. Overall survival was analyzed using multivariable Cox proportional hazards models. To reduce confounding from baseline differences, propensity score matching (PSM) methods were employed in a 1:5 ratio between COH and SEER, respectively, using age group at diagnosis category, gender, race, ethnicity, stage (for solid tumors), and diagnosis year.

**Findings:** Patients treated at COH had significantly superior overall survival (OS) compared with those in the SEER cohort across all examined cancer types after multivariable adjustment for key baseline characteristics and in the PSM analyses. Median OS (when reached) was uniformly longer in the COH cohort compared with the SEER cohort in the full population adjusted model and in the PSM analyses. For the PSM analyses: non-small cell lung cancer (NSCLC) hazard ratio (HR) 0.73 (95% confidence interval [CI]: 0.70-0.76), breast cancer HR 0.83 (95% CI: 0.78-0.86), prostate cancer HR 0.62 (95% CI: 0.58-0.65), colorectal cancer HR 0.74 (95% CI: 0.69-0.79), pancreatic cancer HR 0.75 (95% CI: 0.70-0.81), acute myeloid leukemia HR 0.79 (95% CI: 0.75-0.84), acute lymphoid leukemia HR 0.7 (95% CI: 0.69-0.87), and multiple myeloma HR 0.70 (95% CI: 0.66-0.76) (all *p*<0.001). In addition, notable findings included substantially lower mortality risk for patients with any of the studied advanced solid cancers, including stage IV NSCLC (HR 0.71, 95% CI: 0.67-0.75, *p*<0.001) and stage IV breast cancer (HR 0.78, 95% CI: 0.69-0.87, *p*<0.001).

**Conclusions:** Our results revealed patient care at an individual NCI-CCC employing state-of-the-art therapies and modern care is associated with survival benefits for cancer patients. Patients had significantly superior OS in the COH cohort compared with the SEER cohort for most of the analyzed solid tumors assessed for each stage (I–IV) and for 3 major hematologic malignancies assessed by age group.

Until randomized comparisons studies are possible and the SEER data becomes more comprehensive, the specific causes of the observed benefit and identifying patients that benefit most may be subject to considerable debate. Additional research can help determine with greater granularity the drivers of the survival benefit and highlight an opportunity to make adjusted survival outcomes more accessible to patients, complementing the data currently available because of price transparency mandates. Such efforts may help ensure that all patients benefit from precision medicine approaches.

## Introduction

In 1971, the National Cancer Act established the National Cancer Institute (NCI) Cancer Centers Program; today across the U.S., there are 73 NCI-Designated Cancer Centers, including 57 Comprehensive Cancer Centers (CCC).^1^ These facilities offer a comprehensive approach to cancer prevention, diagnosis, and treatment through tertiary care services such as specialized imaging, complex surgeries, and innovative therapies and technologies available through interventional trials. Tertiary centers are equipped with advanced diagnostic and treatment options not typically available at community hospitals or clinics and are staffed by multidisciplinary teams of surgeons, oncologists, radiologists, pathologists, genetic counselors, pharmacists, social workers, nurses, pain management specialists, and other specialists providing supportive care services. Their coordinated treatment approach facilitates holistic, personalized care.^2^

Tertiary centers serve as regional hubs for sharing expertise and resources with community practices, and extend clinical trial access and specialized care to patients in rural or underserved areas with rare cancers or whose cases require complex management^3–7^. By implementing evidence-based cancer prevention, screening, and treatment strategies tailored to local patient populations, these centers also address cancer disparities in their catchment areas^8^. Through clinical and translational research as well as training programs, they often lead in developing new therapies and care standards and in educating physicians and scientists.

Cancer remains the second leading cause of death in the U.S., where approximately 2 million new diagnoses and over 618,000 deaths are expected in 2025, excluding non-melanoma skin cancers, despite overall reductions in mortality. Reduced tobacco use, earlier detection, new treatments, and gains in access to cancer care, screening, and therapy are among the factors that led to a 34% decline in mortality from 1991 to 2022^9^ and increased rates of 5-year survival from 49% in the 1970s to 69% in the 2010s overall.^10^ However, prognoses still vary widely depending on cancer type and stage at diagnosis as cancers are highly complex and heterogeneous. For instance, while localized breast or prostate cancers are often curable, other advanced-stage cancers have a poorer outlook.

The NCI-CCC model is driven by clinical practice specialization and multidisciplinary, disease-focused care teams, offering expertise informed by clinical trials and biomarker research, as well as treatment regimens tailored to a cancer’s tissue of origin and molecular context.^11–13^ Studies suggest that this model can improve communication, patient satisfaction, and clinical outcomes.^2,14^ Furthermore, evidence from a U.S. study suggests that patients may be underserved in some care settings such that they do not receive appropriate biomarker testing or, even among those who do, the correct treatment.^15^ In that study, 49.7% of patients with non-small cell lung cancer (NSCLC) did not receive biomarker testing results and, of the 50.3% who received testing results with actionable mutations, only 70.8% received the appropriate therapy.^15^ Cancer Centers are often earlier and more efficient adopters of modern approaches to precision oncology.

To evaluate the cancer care and survival outcomes at an NCI-CCC and in the population of the U.S. Surveillance, Epidemiology, and End Results (SEER) national database since 2004, we compared outcomes at our institution (City of Hope National Medical Center [COH]) with the data reported in SEER. Through subspecialized expertise and coordinated care delivery across our network of regional clinical sites, our model supports individualized, biomarker-driven treatment decisions aligned with clinical guidelines (which emerge from coordinated data, shared expertise, and decision support tools^7^) and allows for integration of clinical research into routine practice through timely referral to clinical trials.^16^ Our data suggest that care at an individual Cancer Center employing state-of-the-art, evidence-based therapies and multi-disciplinary care may be associated with survival benefits for cancer.

Currently, there is a lack of transparency and no industry standard for publicly reporting adjusted patient outcomes data in oncology, which makes it difficult for patients and families to find the highest value when seeking cancer care. However, it is already customary for centers to report outcomes for bone marrow transplantation procedures. This study report patient and tumor adjusted models for reporting survival outcomes that could be used as part of a collaborative, transparent model for all cancers.

## Methods

This study investigated the potential advantage associated with receiving cancer care at a high-volume, specialized NCI-CCC (COH). Survival outcomes of patients treated at COH were compared with outcomes reported in the SEER national database. The SEER database collects incident cancer and survival data from national population-based cancer registries and covers approximately 46% of the U.S. population. The SEER registries collect data on patient demographics, primary tumor site, tumor morphology, stage at diagnosis, and patient vital status. This study was approved by the COH institutional review board (IRB 25219).

Patients diagnosed with cancer from 2004 to 2020 were identified from SEER and the COH Cancer Registry (CR). Patient-level SEER data were abstracted using SEER*Stat software, case listing option, from the SEER Research Data (17 registries, November 2023 submission).^17^ All patients queried from the COH CR had to have an abstraction status of complete and coded as an analytic case, as defined by the North American Association of Central Cancer Registries.^18^ Malignant tumors of the following common primary cancer sites and hematologic malignancies with concordant data availability were included: breast, colorectal, lung (non-small cell), prostate, pancreas, multiple myeloma, acute myeloid leukemia (AML), and acute lymphoid leukemia (ALL). The final cohort included patients with the following demographic, clinical, and survival characteristics available in both databases: age at diagnosis, gender, race, ethnicity, primary tumor site, American Joint Committee on Cancer (AJCC) stage (for solid tumors), cancer histologic type, diagnosis year, vital status, and survival time.

The primary endpoint was overall survival (OS), defined from the date of diagnosis to the date of death from any cause or last contact. A study follow-up cut-off date of 12/31/2021 was used to align with the SEER data.

Baseline demographic and clinical characteristics by cancer type were summarized descriptively for each cohort. Multivariable adjustment for age group at diagnosis category (15-44, 45-54, 55-64, 65-74, 75+), gender, race, ethnicity, AJCC stage (for solid tumors), and diagnosis year was performed. In addition, to further reduce confounding from baseline differences and provide more representative data for survival graphs, propensity score matching (PSM) methods^19,20^ were employed using a 1:5 ratio between COH and SEER patients based on those variables, while retaining the adjustment covariables in the Cox regression model. The matching was performed using nearest neighbor methodology without replacement. OS curves comparing COH to SEER were generated by primary cancer type using the Kaplan-Meier method.^21^ Multivariable Cox proportional hazards regression methods comparing COH to SEER were performed separately for each primary cancer type. Adjusted hazard ratios (HRs) and 95% confidence intervals (CIs) were reported from the Cox models.

All reported *P*-values are two-sided. Analyses were performed using R Version 4.4.3.

## Results

### Patient Characteristics

The baseline characteristics of patients in the COH and SEER cohorts are summarized in **Table S1**. A total of 27,394 patients from COH and 3,043,009 patients from SEER were included in the analysis across the eight included cancer types. Compared with the SEER cohort, the COH cohort was younger (median: 61.9 vs. 66.0 years), with a higher proportion in the 15–44 and 45–54 age groups and a lower proportion in the 75+ age group. The COH cohort also had a higher proportion of male patients (54.3% vs. 49.7%), Asian or Pacific Islander patients (15.5% vs. 7.4%), and Hispanic patients (18.6% vs. 10.1%); a lower proportion of Black or African American patients (6.0% vs. 12.0%); and a similar proportion of white patients (77.5% vs. 79.4%). In both cohorts, approximately 66% of patients were diagnosed in 2010 or later. The COH cohort had a higher proportion of patients diagnosed at stage IV for NSCLC (50.5% vs. 43.8%), breast cancer (7.1% vs. 5.2%), and colorectal cancer (38.4% vs. 20.1%), whereas the SEER cohort had a higher proportion of patients diagnosed at stage IV for prostate cancer (9.1% vs. 7.8%) and pancreatic cancer (50.3% vs. 48.6%).

Patients in the COH and SEER cohorts were matched in a 1:5 ratio using propensity scores based on key baseline variables, as described above. After matching, the two cohorts were well balanced across demographic and clinical characteristics, supporting fair comparison of clinical outcomes between populations across diverse cancer types (**Table 1**).

**Table 1.** Demographic and Clinical Characteristics of Patients in the Propensity Score-Matched Cohort.

| Characteristics | NSCLC |  | BREAST |  | PROSTATE |  | COLORECTAL |  | PANCREAS |  | ACUTE MYELOID LEUKEMIA |  | ACUTE LYMPHOID LEUKEMIA |  | MULTIPLE MYELOMA |  |
| --- | --- | --- | --- | --- | --- | --- | --- | --- | --- | --- | --- | --- | --- | --- | --- | --- |
|  | COH<br>(N=2884) | SEER<br>(N=14420) | COH<br>(N=7180) | SEER<br>(N=35900) | COH<br>(N=8961) | SEER<br>(N=44805) | COH<br>(N=2459) | SEER<br>(N=12295) | COH<br>(N=958) | SEER<br>(N=4790) | COH<br>(N=1935) | SEER<br>(N=9675) | COH<br>(N=733) | SEER<br>(N=3665) | COH<br>(N=2284) | SEER<br>(N=11420) |
| **Age, years** |  |  |  |  |  |  |  |  |  |  |  |  |  |  |  |  |
| Mean (SD) | 67.7 (11.5) | 68.6 (11.7) | 57.1 (12.8) | 57.8 (13.1) | 64.0 (8.14) | 64.8 (8.48) | 61.2 (13.6) | 62.1 (14.0) | 66.0 (11.8) | 66.8 (11.6) | 56.2 (16.0) | 57.0 (16.7) | 44.7 (15.8) | 44.8 (16.3) | 59.4 (9.40) | 60.5 (9.81) |
| Median [Min, Max] | 68.6 [22.3, 95.8] | 69.0 [17.0, 90.0] | 56.8 [19.0, 96.9] | 57.0 [17.0, 90.0] | 64.3 [36.8, 94.8] | 65.0 [30.0, 90.0] | 60.9 [17.6, 98.3] | 62.0 [15.0, 90.0] | 66.2 [18.9, 93.2] | 67.0 [15.0, 90.0] | 58.8 [15.2, 94.8] | 60.0 [15.0, 90.0] | 44.6 [20.0, 89.2] | 44.0 [20.0, 90.0] | 60.3 [24.1, 100] | 61.0 [18.0, 90.0] |
| **Age groups, years** |  |  |  |  |  |  |  |  |  |  |  |  |  |  |  |  |
| 15-44 | 93 (3.2%) | 385 (2.7%) | 1296 (18.1%) | 5696 (15.9%) | 78 (0.9%) | 275 (0.6%) | 278 (11.3%) | 1254 (10.2%) | 35 (3.7%) | 159 (3.3%) | 416 (21.5%) | 2061 (21.3%) | 357 (48.7%) | 1834 (50.0%) | 131 (5.7%) | 623 (5.5%) |
| 45-54 | 309 (10.7%) | 1410 (9.8%) | 1947 (27.1%) | 9347 (26.0%) | 1156 (12.9%) | 4695 (10.5%) | 543 (22.1%) | 2551 (20.7%) | 122 (12.7%) | 504 (10.5%) | 323 (16.7%) | 1595 (16.5%) | 146 (19.9%) | 692 (18.9%) | 476 (20.8%) | 2397 (21.0%) |
| 55-64 | 671 (23.3%) | 3133 (21.7%) | 2038 (28.4%) | 10327 (28.8%) | 3580 (40.0%) | 16822 (37.5%) | 633 (25.7%) | 3154 (25.7%) | 271 (28.3%) | 1290 (26.9%) | 506 (26.1%) | 2554 (26.4%) | 137 (18.7%) | 678 (18.5%) | 904 (39.6%) | 4529 (39.7%) |
| 65-74 | 991 (34.4%) | 4950 (34.3%) | 1251 (17.4%) | 6851 (19.1%) | 3372 (37.6%) | 18156 (40.5%) | 596 (24.2%) | 3066 (24.9%) | 316 (33.0%) | 1630 (34.0%) | 469 (24.2%) | 2364 (24.4%) | 72 (9.8%) | 360 (9.8%) | 687 (30.1%) | 3440 (30.1%) |
| 75+ | 820 (28.4%) | 4542 (31.5%) | 648 (9.0%) | 3679 (10.2%) | 775 (8.6%) | 4857 (10.8%) | 409 (16.6%) | 2270 (18.5%) | 214 (22.3%) | 1207 (25.2%) | 221 (11.4%) | 1101 (11.4%) | 21 (2.9%) | 101 (2.8%) | 86 (3.8%) | 431 (3.8%) |
| **Gender** |  |  |  |  |  |  |  |  |  |  |  |  |  |  |  |  |
| Female | 1484 (51.5%) | 7421 (51.5%) | 7180 (100%) | 35900 (100%) | NA | NA | 1149 (46.7%) | 5748 (46.8%) | 449 (46.9%) | 2230 (46.6%) | 898 (46.4%) | 4529 (46.8%) | 343 (46.8%) | 1724 (47.0%) | 1004 (44.0%) | 5032 (44.1%) |
| Male | 1400 (48.5%) | 6999 (48.5%) | NA | NA | 8961 (100%) | 44805 (100%) | 1310 (53.3%) | 6547 (53.2%) | 509 (53.1%) | 2560 (53.4%) | 1037 (53.6%) | 5146 (53.2%) | 390 (53.2%) | 1941 (53.0%) | 1280 (56.0%) | 6388 (55.9%) |
| **Race** |  |  |  |  |  |  |  |  |  |  |  |  |  |  |  |  |
| White | 2058 (71.4%) | 10301 (71.4%) | 5289 (73.7%) | 26425 (73.6%) | 7558 (84.3%) | 37798 (84.4%) | 1790 (72.8%) | 8962 (72.9%) | 715 (74.6%) | 3590 (74.9%) | 1505 (77.8%) | 7584 (78.4%) | 616 (84.0%) | 3147 (85.9%) | 1707 (74.7%) | 8540 (74.8%) |
| Black or African American | 120 (4.2%) | 598 (4.1%) | 411 (5.7%) | 2045 (5.7%) | 512 (5.7%) | 2557 (5.7%) | 119 (4.8%) | 589 (4.8%) | 59 (6.2%) | 298 (6.2%) | 80 (4.1%) | 397 (4.1%) | 19 (2.6%) | 92 (2.5%) | 314 (13.7%) | 1569 (13.7%) |
| Asian or Pacific Islander | 685 (23.8%) | 3437 (23.8%) | 1393 (19.4%) | 7003 (19.5%) | 824 (9.2%) | 4115 (9.2%) | 521 (21.2%) | 2622 (21.3%) | 174 (18.2%) | 872 (18.2%) | 325 (16.8%) | 1624 (16.8%) | 88 (12.0%) | 401 (10.9%) | 245 (10.7%) | 1229 (10.8%) |
| Others/Unknown | 21 (0.7%) | 84 (0.6%) | 87 (1.2%) | 427 (1.2%) | 67 (0.7%) | 335 (0.7%) | 29 (1.2%) | 122 (1.0%) | 10 (1.0%) | 30 (0.6%) | 25 (1.3%) | 70 (0.7%) | 10 (1.4%) | 25 (0.7%) | 18 (0.8%) | 82 (0.7%) |
| **Ethnicity** |  |  |  |  |  |  |  |  |  |  |  |  |  |  |  |  |
| Non-hispanic | 2593 (89.9%) | 12967 (89.9%) | 5254 (73.2%) | 26251 (73.1%) | 8078 (90.1%) | 40396 (90.2%) | 1938 (78.8%) | 9709 (79.0%) | 777 (81.1%) | 3874 (80.9%) | 1558 (80.5%) | 7777 (80.4%) | 386 (52.7%) | 1912 (52.2%) | 1708 (74.8%) | 8544 (74.8%) |
| Hispanic | 291 (10.1%) | 1453 (10.1%) | 1926 (26.8%) | 9649 (26.9%) | 883 (9.9%) | 4409 (9.8%) | 521 (21.2%) | 2586 (21.0%) | 181 (18.9%) | 916 (19.1%) | 377 (19.5%) | 1898 (19.6%) | 347 (47.3%) | 1753 (47.8%) | 576 (25.2%) | 2876 (25.2%) |
| **Stage** |  |  |  |  |  |  |  |  |  |  |  |  |  |  |  |  |
| 0 | NA | NA | 39 (0.5%) | 137 (0.4%) | NA | NA | 22 (0.9%) | 113 (0.9%) | NA | NA | NA | NA | NA | NA | NA | NA |
| I | 611 (21.2%) | 3067 (21.3%) | 2922 (40.7%) | 14614 (40.7%) | 402 (4.5%) | 2000 (4.5%) | 345 (14.0%) | 1727 (14.0%) | 105 (11.0%) | 530 (11.1%) | NA | NA | NA | NA | NA | NA |
| II | 210 (7.3%) | 1052 (7.3%) | 2642 (36.8%) | 13227 (36.8%) | 6478 (72.3%) | 32401 (72.3%) | 409 (16.6%) | 2047 (16.6%) | 233 (24.3%) | 1169 (24.4%) | NA | NA | NA | NA | NA | NA |
| III | 607 (21.0%) | 3024 (21.0%) | 1066 (14.8%) | 5356 (14.9%) | 1381 (15.4%) | 6900 (15.4%) | 738 (30.0%) | 3692 (30.0%) | 154 (16.1%) | 762 (15.9%) | NA | NA | NA | NA | NA | NA |
| IV | 1456 (50.5%) | 7277 (50.5%) | 511 (7.1%) | 2566 (7.1%) | 700 (7.8%) | 3504 (7.8%) | 945 (38.4%) | 4716 (38.4%) | 466 (48.6%) | 2329 (48.6%) | NA | NA | NA | NA | NA | NA |
| **Diagnosis year** |  |  |  |  |  |  |  |  |  |  |  |  |  |  |  |  |
| <2010 | 810 (28.1%) | 4059 (28.1%) | 1839 (25.6%) | 9173 (25.6%) | 4490 (50.1%) | 22443 (50.1%) | 718 (29.2%) | 3595 (29.2%) | 202 (21.1%) | 1012 (21.1%) | 533 (27.5%) | 2660 (27.5%) | 168 (22.9%) | 837 (22.8%) | 469 (20.5%) | 2347 (20.6%) |
| ≥2010 | 2074 (71.9%) | 10361 (71.9%) | 5341 (74.4%) | 26727 (74.4%) | 4471 (49.9%) | 22362 (49.9%) | 1741 (70.8%) | 8700 (70.8%) | 756 (78.9%) | 3778 (78.9%) | 1402 (72.5%) | 7015 (72.5%) | 565 (77.1%) | 2828 (77.2%) | 1815 (79.5%) | 9073 (79.4%) |

### Survival Outcomes by Cancer Type and Stratified

Patients treated at COH had significantly longer OS compared with those in the SEER cohort across all examined cancer types, using a multivariable model with adjustment for key baseline characteristics in both the original and PSM populations. Median OS (when reached) was uniformly longer in the COH cohort compared with the SEER cohort in the PSM analyses. For NSCLC, median OS was 2.20 years (95% CI: 2.05–2.33) at COH versus 1.42 years (95% CI: 1.42–1.50) in SEER, corresponding to an adjusted HR of 0.73 (95% CI: 0.70–0.76). For colorectal cancer, median OS was 7.31 years (95% CI: 6.57–8.16) at COH versus 5.25 years (95% CI: 5.00–5.58) in SEER, with adjusted HR 0.74 (95% CI: 0.69–0.79). For pancreatic cancer, median OS was 1.14 years (95% CI: 1.06–1.22) at COH versus 0.75 years (95% CI: 0.67–0.75) in SEER, with adjusted HR 0.75 (95% CI: 0.70–0.81). For AML, median OS was 2.37 years (95% CI: 2.09–2.81) at COH versus 1.50 years (95% CI: 1.42–1.58) in SEER, with adjusted HR 0.79 (95% CI: 0.75–0.84). For ALL, median OS was 6.36 years (95% CI: 4.78–13.45) at COH versus 3.67 years (95% CI: 3.25–4.08) in SEER, with adjusted HR 0.77 (95% CI: 0.69–0.87). For multiple myeloma, median OS was 9.40 years (95% CI: 8.81–10.05) at COH versus 7.17 years (95% CI: 6.92–7.42) in SEER, with adjusted HR 0.70 (95% CI: 0.66–0.76). For breast and prostate cancer, the median OS was not reached (NR) in either cohort; however, COH patients had improved outcomes compared with SEER: for breast cancer, 5-year survival was 87% versus 84% with an adjusted HR of 0.83 (95% CI: 0.79–0.86); for prostate cancer 5-year survival was 93% versus 89% with an adjusted HR of 0.62 (95% CI: 0.58–0.65). Across all cancer types, the direction and magnitude of benefit favored COH, with p<0.001 for all comparisons (**Figure 1**, **Table 2**). Comparisons of the unmatched cohorts after multivariable adjustment for key baseline characteristics are provided in the Supplement (**Figure S1**, **Table 2**). The direction and significance of the survival benefit was consistent across the original population and PSM population, supporting the robustness of the findings.

**Figure 1.**
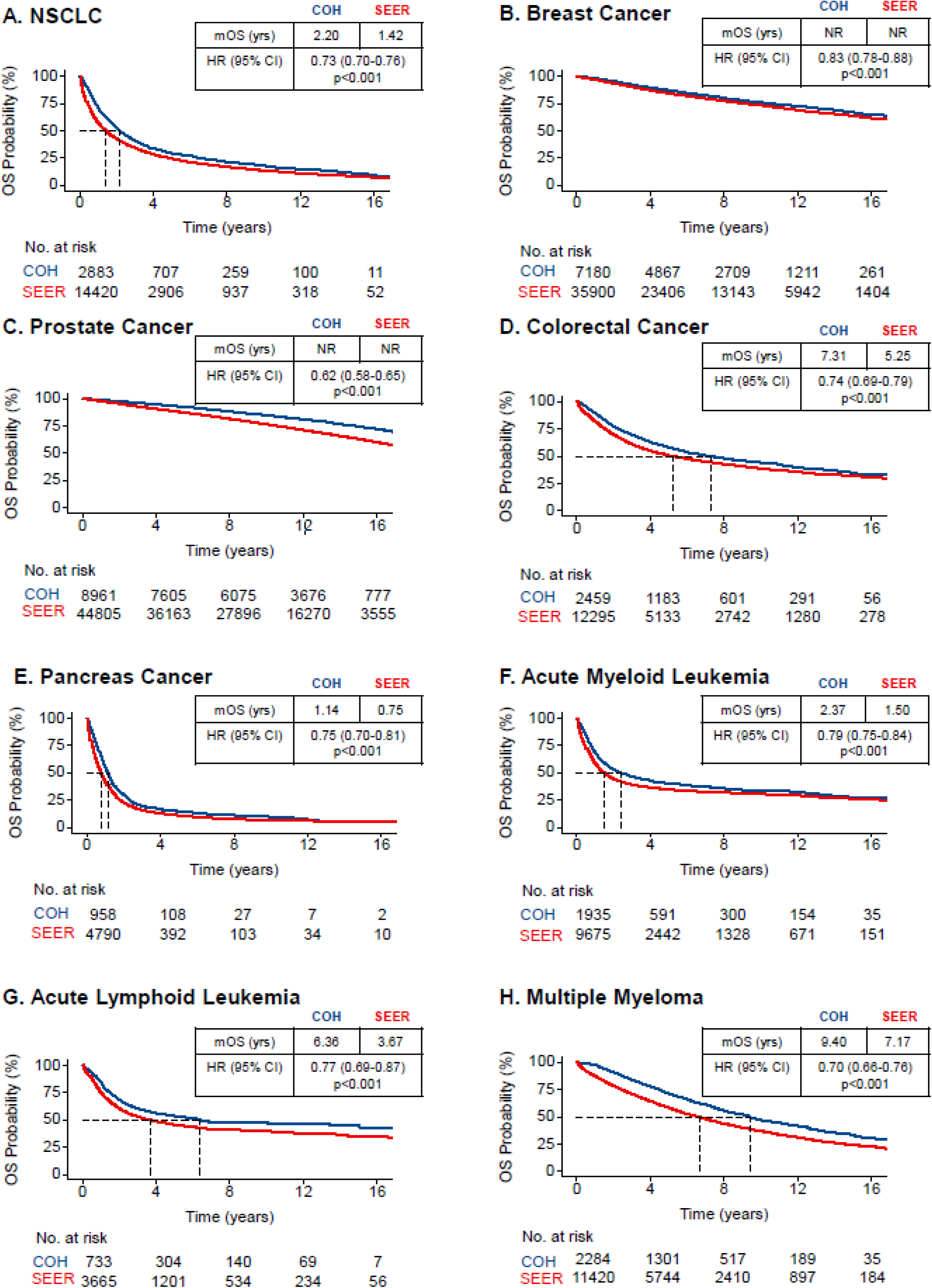
Overall survival comparisons between propensity score-matched City of Hope and SEER cohorts by Kaplan-Meier analysis for 8 cancer types. (A) NSCLC. (B) Breast cancer. (C) Prostate cancer. (D) Colorectal cancer. (E) Pancreas cancer. (F) Acute myeloid leukemia. (G) Acute lymphoid leukemia. (H) Multiple myeloma. CI, confidence interval; HR, hazard ratio; mOS, median overall survival; NR, not reached; NSCLC, non-small cell lung cancer; OS, overall survival.

**Table 2.** Comparison of Median Overall Survival and Hazard Ratios Between City of Hope and SEER Cohorts Across Cancer Types.

| Cancer type | Cohort | Based on original population |  | Based on PSM population |  |
| --- | --- | --- | --- | --- | --- |
|  |  | Median OS (years), 95% CI | Hazard ratio (COH vs SEER) | Median OS (years), 95% CI | Hazard ratio (COH vs SEER) |
| NSCLC | COH | 2.20 (2.05, 2.33) | 0.67 (0.64-0.70, p<0.001) | 2.20 (2.05, 2.33) | 0.73 (0.70-0.76, p<0.001) |
|  | SEER | 1.25 (1.25, 1.25) | ref | 1.42 (1.42, 1.50) | ref |
| breast cancer | COH | NR | 0.78 (0.74-0.82, p<0.001) | NR | 0.83 (0.78-0.88, p<0.001) |
|  | SEER | NR | ref | NR | ref |
| prostate cancer | COH | NR | 0.55 (0.53-0.58, p<0.001) | NR | 0.62 (0.58-0.65, p<0.001) |
|  | SEER | 16 (15.9, 16.0) | ref | NR | ref |
| colorectal cancer | COH | 7.31 (6.57, 8.16) | 0.72 (0.68-0.76, p<0.001) | 7.31 (6.57, 8.16) | 0.74 (0.69-0.79, p<0.001) |
|  | SEER | 7.17 (7.08, 7.17) | ref | 5.25 (5.00, 5.58) | ref |
| pancreatic cancer | COH | 1.14 (1.06, 1.22) | 0.71 (0.66-0.76, p<0.001) | 1.14 (1.06, 1.22) | 0.75 (0.70-0.81, p<0.001) |
|  | SEER | 0.67 (0.67, 0.67) | ref | 0.75 (0.67, 0.75) | ref |
| acute myeloid leukemia | COH | 2.37 (2.09, 2.81) | 0.74 (0.70-0.79, p<0.001) | 2.37 (2.09, 2.81) | 0.79 (0.75-0.84, p<0.001) |
|  | SEER | 1.00 (1.00, 1.00) | ref | 1.50 (1.42, 1.58) | ref |
| acute lymphoid leukemia | COH | 6.36 (4.78, 13.45) | 0.71 (0.63-0.79, p<0.001) | 6.36 (4.78, 13.4) | 0.77 (0.69-0.87, p<0.001) |
|  | SEER | 2.42 (2.25, 2.58) | ref | 3.67 (3.25, 4.08) | ref |
| multiple myeloma | COH | 9.4 (8.81, 10.05) | 0.65 (0.61-0.70, p<0.001) | 9.40 (8.81, 10.05) | 0.70 (0.66-0.76, p<0.001) |
|  | SEER | 4.5 (4.5, 4.58) | ref | 7.17 (6.92, 7.42) | ref |

NR, not reached. NE, not estimable. CI, confidence interval. OS, overall survival. PSM, propensity score-matching. Ref, reference. All Hazard Ratios are adjusted for age category, gender (except for breast and prostate cancer), race, ethnicity, AJCC stage (for solid tumors), and diagnosis year. PSM was propensity score matched, with the Hazard Ratio estimated in a model that retained adjustment for those key characteristics.

Similarly, patients in the COH cohort had significantly longer OS compared with those in the SEER cohort when analyzed by various stages of diagnosis for solid tumors (stage I–IV) or by age for hematologic malignancies (age <60 years or ≥60 years for AML and ALL; age <60 years, 60–70 years, and >70 years for multiple myeloma) (**Figures 2–3**, **S2–S7**; **Tables S2–S3**). Hazard ratios demonstrating lower mortality risk not reaching significance included stage I prostate cancer (median OS not reached in COH cohort, 15 years in SEER cohort), stage I and II colorectal cancer (stage I median OS not reached in COH cohort, 16.8 years in SEER cohort; stage II median OS 14.8 vs. 13.2 years), stage I pancreatic cancer (median OS 3.77 vs 2.08 years), AML in patients <60 years (median OS 6.1 vs. 7.8 years), and ALL in patients ≥60 years (median OS 2.02 vs. 1.67 years). PSM hazard ratio analysis demonstrated that patients in the COH cohort diagnosed with stage IV solid tumors had lower morality risk compared with the respective SEER patients: NSCLC HR 0.71 (95% CI: 0.67–0.75), breast cancer HR 0.78 (95% CI: 0.69–0.87), prostate cancer HR 0.58 (95% CI: 0.52–0.66), colorectal cancer HR 0.68 (95% CI: 0.62–0.74), and pancreatic cancer HR 0.74 (95% CI: 0.66–0.82) (all *p*<0.001).

**Figure 2.**
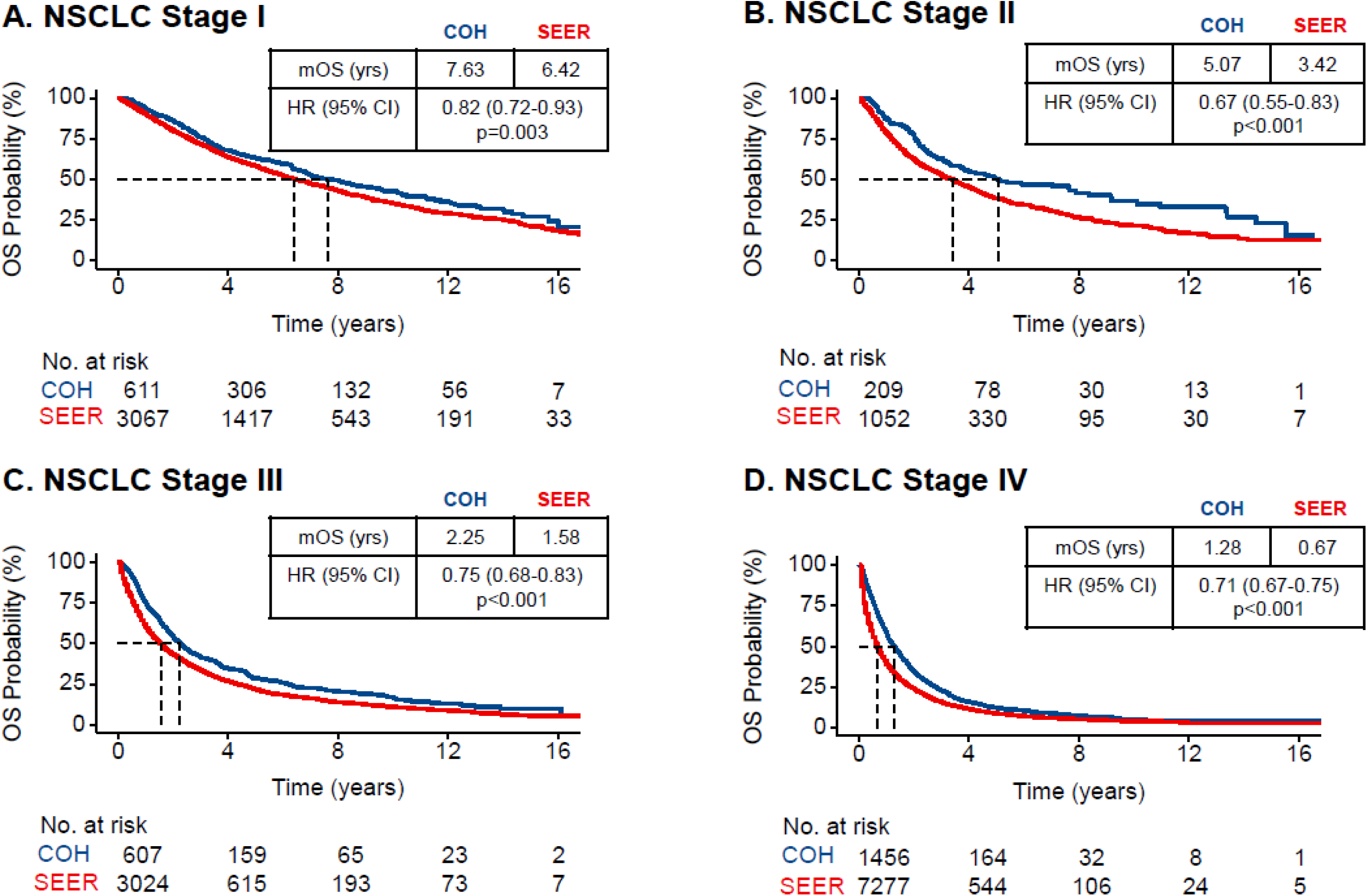
Overall survival comparisons between propensity score-matched City of Hope and SEER cohorts by Kaplan-Meier analysis for NSCLC stages I–IV. (A) Patients with Stage I; (B) Patients with Stage II; (C) Patients with Stage III; (D) Patients with Stage IV. CI, confidence interval; HR, hazard ratio; mOS, median overall survival; NR, not reached; OS, overall survival.

**Figure 3.**
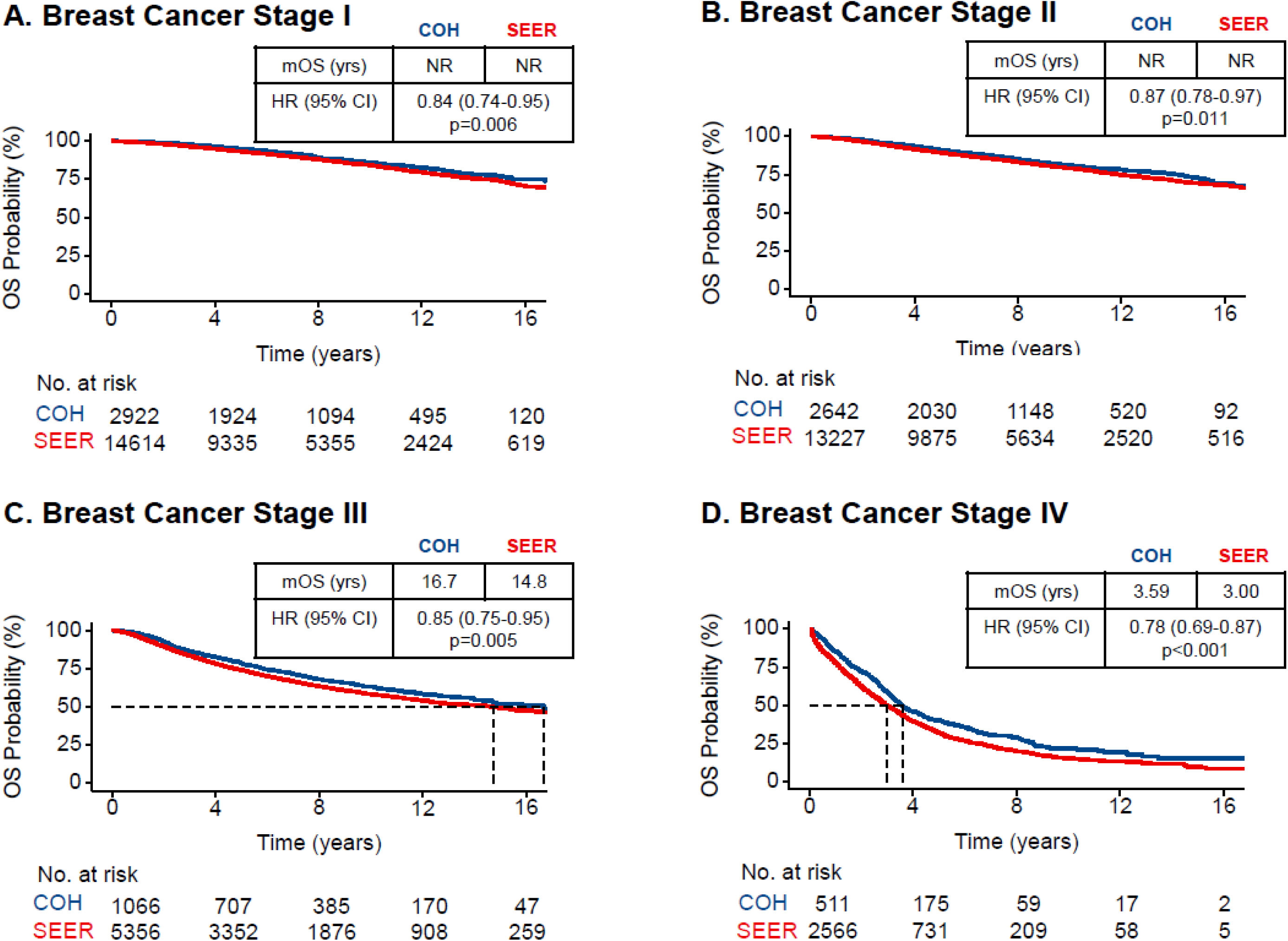
Overall survival comparisons between propensity score-matched City of Hope and SEER cohorts by Kaplan-Meier analysis for breast cancer stages I–IV. (A) Patients with Stage I; (B) Patients with Stage II; (C) Patients with Stage III; (D) Patients with Stage IV. CI, confidence interval; HR, hazard ratio; mOS, median overall survival; NR, not reached; OS, overall survival.

## Discussion

In this study, we found that patients treated at COH had significantly better OS compared to patients with similar cancers in the SEER national cancer registry. This survival advantage was observed across eight cancer types. Notable findings include substantially lower mortality risk at COH for patients with several advanced stage cancers, including stage IV NSCLC (HR 0.71), stage IV breast cancer (HR 0.78), as well as for patients with hematologic malignancies including AML (HR 0.70 for patients aged ≥60 years) and multiple myeloma (HR 0.72 for patients aged ≥70 years). These findings likely reflect several factors associated with care at a specialized cancer center.

Several other studies have reported that care at an NCI-CCC is associated with superior survival. In a systematic review and meta-analysis, Thamm et al. reported superior mortality and survival outcomes, along with other positive outcome measures, for adults with solid cancer treated at an NCI-CCC compared with non-comprehensive treatment site, including 23% lower overall mortality risk^22^. Wolfson et al. found that, among patients with newly diagnosed adult-onset cancers in Los Angeles County between 1998–2008, those first treated at a care setting other than an NCI-CCC had a 20%–50% increased risk of mortality, including for hepatobiliary, lung, pancreatic, gastric, breast, and colorectal cancers^23^. This analysis adjusted for clinical and sociodemographic factors including age at diagnosis, sex, race/ethnicity, stage of disease, socioeconomic status, payor, and distance to the nearest center. In another study, Bruno et al. compared outcomes among U.S. patients treated for small lymphocytic lymphoma/chronic lymphocytic leukemia (CLL) or Mantle cell lymphoma (MCL) between 2013–2022, reporting significantly longer survival in the group treated at an academic center versus a community clinic^24^. This analysis adjusted for diagnosis year, age, sex/race/ethnicity, socioeconomic status, clinical practice size, insurance type, and disease-specific variables.

These studies hypothesize several factors potentially leading to superior outcomes at a tertiary cancer center, including care quality and compliance with treatment guidelines, the availability of clinical trials and newer treatments such as immunotherapies, multidisciplinary disease management and shared records, and systems for integrated supportive care.^23,24^ In 2024, Unger et al. reported that, at NCI-CCC Programs, 21.6% of patients are involved in commercial clinical trials or investigator-initiated therapeutic trials, compared with <6% of patients at other academic centers, integrated network cancer programs, and community practices.^25^ Similarly, COH prioritizes trial enrollment for all patients, which may offer personalized treatment strategies to benefit patients with relapsed, treatment-resistant, and persistent cancers. Notably, despite increased access to clinical trials at COH, our dataset includes all patients treated and therefore the survival rates are expected to be lower than those observed in randomized clinical trials. For instance, recent clinical trials of gemcitabine and abraxane for metastatic pancreatic cancer reported median overall survival of 10 months^26^, whereas the survival rate is lower in our stage IV cohort, which includes patients who would not be eligible for a trial owing to poor performance status or other issues related to frailty or inability to receive therapy.

NCI-CCCs offer multidisciplinary care teams, access to clinical trials, advanced imaging and diagnostics, and individualized treatment planning to deliver therapeutically precise care. Clinical trials at our center alone have yielded groundbreaking discoveries, including novel targeted therapies and immunotherapy regimens, first-in-class CAR T cell therapies, advanced bone marrow transplant regimens, AI-driven early cancer detection, and robotic cancer surgeries^27–31^. Tumor profiling (genomic, proteomic, metabolic, etc.), biomarker testing, and advanced imaging have improved our ability to offer treatments informed by the molecular characteristics of each cancer. Furthermore, molecular tumor boards, pyramidal decision-support structures, integrated data systems, and enterprise-wide diagnostic tools provide the infrastructure supporting precision medicine, and these systems increasingly integrate artificial intelligence (AI)-based decision platforms and drug-matching algorithms.^3,5,32–34^ In our analysis, differences in survival were especially notable in cancers with generally poor prognoses, where expertise and early intervention may have a greater impact on outcomes. Additionally, patient-centered, coordinated care may contribute to improved survival outcomes and quality of life.

Subspecialized clinical practice at an NCI-CCC may also contribute to patient outcomes, as different cancers require distinct treatment modalities. For instance, these centers are among those that can deliver definitive therapy of stem cell transplantation for immune reconstitution or newer immune-based therapies, such as CAR T cell therapy, for relapsed or refractory hematologic diseases^35,36^. For solid tumors, treatment often remains guided by tumor-specific mutations and biology.^37,38^ As of early 2025, there were nine Food and Drug Administration (FDA)-approved tumor-agnostic therapies, compared to hundreds of mutation-targeted and tumor-specific drugs^12,39^ We hypothesize that NCI-CCCs are well equipped to use tumor-agnostic therapies that nonetheless require deep understanding of specific tumor characteristics and genomics.

In this study, we compared patient survival outcomes at a single institution versus the publicly available SEER database. For patients and families who seek optimal outcomes, there is no industry standard for publicly reporting adjusted patient outcomes data that would provide meaningful information to them. While in the domain of blood and marrow transplantation, a government-mandated survival outcome model allows stakeholders to assess centers by their risk-adjusted survival outcomes for patients who undergo allogeneic transplantation, it is seldom used by patients (and not consistently by referring physicians or payers), and no such transparency exists in any other domain of oncology.

While we commend the federal government for the progress made to advance price transparency in recent years, the absence of outcomes data leaves patients and families with a key missing variable in finding the highest value in their cancer care. Our study, therefore, is intended to be a call to action to all cancer centers and cancer care providers to develop transparent, appropriately risk-adjusted models that would allow for public reporting and center accountability for survival outcomes. A successful model for blood and marrow transplant patients already exists, and we believe that it can be a blueprint for the collaborative creation of a similarly transparent model for survival outcomes for all cancers. This paper advances this much-needed process.

### Limitations

Despite adjusting for key demographic and clinical variables, this study has several limitations related to those of the SEER dataset, which lacks important variables affecting cancer survival, including comorbidities, performance status, organ function, socioeconomic factors and geolocation, treatment intent and detailed history (chemotherapy regimen, radiation dose, surgical quality, etc.), and molecular testing results. Therefore, these variables were not included in the analysis and residual confounding is possible. Other national databases such as NCDB are also limited by lack of key data points, including comorbidities, socioeconomic factors, and molecular testing results. Furthermore, known disparities related to cancer treatment and survival outcomes include rural versus urban treatment sites, community hospitals versus academic centers, and different clinical trial access between populations. The SEER dataset includes a heterogeneous cohort of hundreds of thousands of patients across the U.S. treated in differing care settings, whereas the COH cohort reflects the demographics in our Southern California catchment area and includes patients treated at a tertiary center. As Los Angeles and the region contribute data to the SEER program, there may be patient overlap that is unaccounted for that would impact the independence of the estimates, although the proportional contribution of COH data to SEER is small and unlikely to impact the results.

Referral bias is another potential confounder. Patients treated at a center like COH may differ systematically from those treated in community settings who are included in the SEER dataset. For example, patients at a tertiary center may be more likely to travel for care, have better functional status, or be more engaged in the healthcare system, all of which could influence outcomes independently of treatment received. However, there may be a higher number of patients referred to a tertiary center for treatment of more complex or advanced diseases. While we used propensity score matching to balance measured characteristics between the two cohorts, unmeasured factors related to referral patterns could still introduce bias. COH follows the latest AJCC staging guidelines enterprise-wide, whereas SEER staging includes the AJCC staging system current at the time of diagnosis rather than at the time of data entry. SEER also does not capture institutional characteristics or care delivery metrics, which may also play a role in outcomes.

Furthermore, the COH dataset may differ from other NCI-CCCs as it includes a clinical network of coordinated care delivery beyond our academic cancer center. We have prioritized standardized protocols, physician education, and clinical trial infrastructure and implemented routine liquid biopsy, multi-omics analyses, and integrated data systems to match patients to targeted therapies and track treatment response and resistance. Our results point to the benefits of aligning community practices with a cancer center, as this model of high-quality, precision medicine–guided care delivered by integrated, multidisciplinary teams may lead to superior clinical outcomes across diverse cancers.

Despite these limitations, our results were similar in the adjusted and propensity score matched analyses and consistent with prior reports evaluating tertiary cancer centers.

## Conclusion

Overall, our findings suggest that specialized cancer care at an NCI-CCC is associated with longer survival across a range of malignancies. These results support the value of high-volume, multidisciplinary cancer centers and may inform efforts to improve access to specialized care, optimize referral practices, and address disparities in cancer outcomes. Future studies could consider data from other sources such as the National Cancer Database (NCDB) and other cancer centers, which may allow for comparison of the long-term trends in cancer survival in different settings and for analysis of more detailed center, patient, and treatment characteristics. Cancer care has improved substantially in the last decade owing to advances in next-generation sequencing, liquid biopsy, surgery, radiation, and therapeutics, including precision medicine approaches. Price transparency and outcomes data have historically lagged innovation, but multicenter data sharing and collaboration to develop a standardized model to publicly report the adjusted survival outcomes of all cancer centers would provide meaningful data and information to patients and their families. The analysis reported here should spur further research to identify with greater granularity and evidence the drivers of the survival advantages at individual academic centers and in different regions, which may offer insights to help ensure improved outcomes for all patients. Advances in cancer care such as genomics approaches as well as complex and targeted therapies offer great promise but also risk exacerbating disparities in patient outcomes that already exist. Follow-up studies to this analysis should promote the development of better guidelines and care models to improve outcomes for all patients.

## Contributions

R.S., J.A., V.T., and M.V.D.B. conceived, designed, and oversaw the study. The manuscript was originally drafted by R.S., A.V., X.L., J.A.R., J.F., I.M., H.R., and M.V.D.B with feedback from all authors. A.V. and X.L. collected the data, validated the data, performed primary statistical analysis and helped draft the manuscript. P.F. assisted with statistical analysis. A.V., X.L., J.A.R., J.F., P.F., I.M., H.R., and H.L. reviewed data visualization, data presentation, and contributed to the discussion of the study. All authors critically reviewed the manuscript, provided edits and approved the final article and take responsibility for its content.

## Declarations

This study was conducted according to the guidelines of the Declaration of Helsinki and approved by the City of Hope institutional review board (IRB #25219).

## Data Availability

All data produced in the present work are contained in the manuscript.

---

**Supplemental Table 1.** Demographic and Clinical Characteristics of Patients in the Original Cohort.

| Characteristics | NSCLC |  | BREAST |  | PROSTATE |  | COLORECTAL |  | PANCREAS |  | ACUTE MYELOID LEUKEMIA |  | ACUTE LYMPHOID LEUKEMIA |  | MULTIPLE MYELOMA |  |
| --- | --- | --- | --- | --- | --- | --- | --- | --- | --- | --- | --- | --- | --- | --- | --- | --- |
|  | COH<br>(N=2884) | SEER<br>(N=555133) | COH<br>(N=7180) | SEER<br>(N=893557) | COH<br>(N=8961) | SEER<br>(N=810914) | COH<br>(N=2459) | SEER<br>(N=508600) | COH<br>(N=958) | SEER<br>(N=135396) | COH<br>(N=1935) | SEER<br>(N=41651) | COH<br>(N=733) | SEER<br>(N=9488) | COH<br>(N=2284) | SEER<br>(N=88270) |
| **Age, years** |  |  |  |  |  |  |  |  |  |  |  |  |  |  |  |  |
| Mean (SD) | 67.7 (11.5) | 69.3 (10.9) | 57.1 (12.8) | 61.4 (13.6) | 64.0 (8.14) | 66.5 (9.12) | 61.2 (13.6) | 66.1 (13.9) | 66.0 (11.8) | 68.0 (12.0) | 56.2 (16.0) | 62.7 (17.7) | 44.7 (15.8) | 49.7 (18.5) | 59.4 (9.40) | 67.9 (12.0) |
| Median [Min, Max] | 68.6 [22.3, 95.8] | 70.0 [15.0, 90.0] | 56.8 [19.0, 96.9] | 62.0 [15.0, 90.0] | 64.3 [36.8, 94.8] | 66.0 [23.0, 90.0] | 60.9 [17.6, 98.3] | 67.0 [15.0, 90.0] | 66.2 [18.9, 93.2] | 69.0 [15.0, 90.0] | 58.8 [15.2, 94.8] | 66.0 [15.0, 90.0] | 44.6 [20.0, 89.2] | 50.0 [20.0, 90.0] | 60.3 [24.1, 100] | 69.0 [16.0, 90.0] |
| **Age groups, years** |  |  |  |  |  |  |  |  |  |  |  |  |  |  |  |  |
| 15-44 | 93 (3.2%) | 8983 (1.6%) | 1296 (18.1%) | 100405 (11.2%) | 78 (0.9%) | 4031 (0.5%) | 278 (11.3%) | 32142 (6.3%) | 35 (3.7%) | 4218 (3.1%) | 416 (21.5%) | 6835 (16.4%) | 357 (48.7%) | 3882 (40.9%) | 131 (5.7%) | 2851 (3.2%) |
| 45-54 | 309 (10.7%) | 43494 (7.8%) | 1947 (27.1%) | 187590 (21.0%) | 1156 (12.9%) | 71010 (8.8%) | 543 (22.1%) | 76678 (15.1%) | 122 (12.7%) | 13564 (10.0%) | 323 (16.7%) | 4840 (11.6%) | 146 (19.9%) | 1586 (16.7%) | 476 (20.8%) | 9709 (11.0%) |
| 55-64 | 671 (23.3%) | 124423 (22.4%) | 2038 (28.4%) | 229764 (25.7%) | 3580 (40.0%) | 264138 (32.6%) | 633 (25.7%) | 115137 (22.6%) | 271 (28.3%) | 32798 (24.2%) | 506 (26.1%) | 7660 (18.4%) | 137 (18.7%) | 1737 (18.3%) | 904 (39.6%) | 20628 (23.4%) |
| 65-74 | 991 (34.4%) | 188980 (34.0%) | 1251 (17.4%) | 211951 (23.7%) | 3372 (37.6%) | 316799 (39.1%) | 596 (24.2%) | 129108 (25.4%) | 316 (33.0%) | 41952 (31.0%) | 469 (24.2%) | 10261 (24.6%) | 72 (9.8%) | 1316 (13.9%) | 687 (30.1%) | 27044 (30.6%) |
| 75+ | 820 (28.4%) | 189253 (34.1%) | 648 (9.0%) | 163847 (18.3%) | 775 (8.6%) | 154936 (19.1%) | 409 (16.6%) | 155535 (30.6%) | 214 (22.3%) | 42864 (31.7%) | 221 (11.4%) | 12055 (28.9%) | 21 (2.9%) | 967 (10.2%) | 86 (3.8%) | 28038 (31.8%) |
| **Gender** |  |  |  |  |  |  |  |  |  |  |  |  |  |  |  |  |
| Female | 1484 (51.5%) | 264527 (47.7%) | 7180 (100%) | 893557 (100%) | NA | NA | 1149 (46.7%) | 242850 (47.7%) | 449 (46.9%) | 65971 (48.7%) | 898 (46.4%) | 18698 (44.9%) | 343 (46.8%) | 4285 (45.2%) | 1004 (44.0%) | 39228 (44.4%) |
| Male | 1400 (48.5%) | 290606 (52.3%) | NA | NA | 8961 (100%) | 810914 (100%) | 1310 (53.3%) | 265750 (52.3%) | 509 (53.1%) | 69425 (51.3%) | 1037 (53.6%) | 22953 (55.1%) | 390 (53.2%) | 5203 (54.8%) | 1280 (56.0%) | 49042 (55.6%) |
| **Race** |  |  |  |  |  |  |  |  |  |  |  |  |  |  |  |  |
| White | 2058 (71.4%) | 451225 (81.3%) | 5289 (73.7%) | 712652 (79.8%) | 7558 (84.3%) | 636480 (78.5%) | 1790 (72.8%) | 402006 (79.0%) | 715 (74.6%) | 108081 (79.8%) | 1505 (77.8%) | 33796 (81.1%) | 616 (84.0%) | 7818 (82.4%) | 1707 (74.7%) | 64484 (73.1%) |
| Black or African American | 120 (4.2%) | 60149 (10.8%) | 411 (5.7%) | 92105 (10.3%) | 512 (5.7%) | 117994 (14.6%) | 119 (4.8%) | 56578 (11.1%) | 59 (6.2%) | 15243 (11.3%) | 80 (4.1%) | 3639 (8.7%) | 19 (2.6%) | 676 (7.1%) | 314 (13.7%) | 17362 (19.7%) |
| Asian or Pacific Islander | 685 (23.8%) | 39820 (7.2%) | 1393 (19.4%) | 78999 (8.8%) | 824 (9.2%) | 41427 (5.1%) | 521 (21.2%) | 43261 (8.5%) | 174 (18.2%) | 10909 (8.1%) | 325 (16.8%) | 3784 (9.1%) | 88 (12.0%) | 819 (8.6%) | 245 (10.7%) | 5270 (6.0%) |
| Others/Unknown | 21 (0.7%) | 3939 (0.7%) | 87 (1.2%) | 9801 (1.1%) | 67 (0.7%) | 15013 (1.9%) | 29 (1.2%) | 6755 (1.3%) | 10 (1.0%) | 1163 (0.9%) | 25 (1.3%) | 432 (1.0%) | 10 (1.4%) | 175 (1.8%) | 18 (0.8%) | 1154 (1.3%) |
| **Ethnicity** |  |  |  |  |  |  |  |  |  |  |  |  |  |  |  |  |
| Non-hispanic | 2593 (89.9%) | 520976 (93.8%) | 5254 (73.2%) | 791478 (88.6%) | 8078 (90.1%) | 734376 (90.6%) | 1938 (78.8%) | 450117 (88.5%) | 777 (81.1%) | 120023 (88.6%) | 1558 (80.5%) | 36131 (86.7%) | 386 (52.7%) | 6289 (66.3%) | 1708 (74.8%) | 77637 (88.0%) |
| Hispanic | 291 (10.1%) | 34157 (6.2%) | 1926 (26.8%) | 102079 (11.4%) | 883 (9.9%) | 76538 (9.4%) | 521 (21.2%) | 58483 (11.5%) | 181 (18.9%) | 15373 (11.4%) | 377 (19.5%) | 5520 (13.3%) | 347 (47.3%) | 3199 (33.7%) | 576 (25.2%) | 10633 (12.0%) |
| **Stage** |  |  |  |  |  |  |  |  |  |  |  |  |  |  |  |  |
| 0 | NA | NA | 39 (0.5%) | 1393 (0.2%) | NA | NA | 22 (0.9%) | 17267 (3.4%) | NA | NA | NA | NA | NA | NA | NA | NA |
| I | 611 (21.2%) | 146553 (26.4%) | 2922 (40.7%) | 474828 (53.1%) | 402 (4.5%) | 128631 (15.9%) | 345 (14.0%) | 126937 (25.0%) | 105 (11.0%) | 17827 (13.2%) | NA | NA | NA | NA | NA | NA |
| II | 210 (7.3%) | 43971 (7.9%) | 2642 (36.8%) | 274009 (30.7%) | 6478 (72.3%) | 519537 (64.1%) | 409 (16.6%) | 127771 (25.1%) | 233 (24.3%) | 34661 (25.6%) | NA | NA | NA | NA | NA | NA |
| III | 607 (21.0%) | 121503 (21.9%) | 1066 (14.8%) | 96973 (10.9%) | 1381 (15.4%) | 89197 (11.0%) | 738 (30.0%) | 134405 (26.4%) | 154 (16.1%) | 14834 (11.0%) | NA | NA | NA | NA | NA | NA |
| IV | 1456 (50.5%) | 243106 (43.8%) | 511 (7.1%) | 46354 (5.2%) | 700 (7.8%) | 73549 (9.1%) | 945 (38.4%) | 102220 (20.1%) | 466 (48.6%) | 68074 (50.3%) | NA | NA | NA | NA | NA | NA |
| **Diagnosis year** |  |  |  |  |  |  |  |  |  |  |  |  |  |  |  |  |
| <2010 | 810 (28.1%) | 187238 (33.7%) | 1839 (25.6%) | 288096 (32.2%) | 4490 (50.1%) | 301471 (37.2%) | 718 (29.2%) | 181714 (35.7%) | 202 (21.1%) | 36956 (27.3%) | 533 (27.5%) | 11820 (28.4%) | 168 (22.9%) | 2787 (29.4%) | 469 (20.5%) | 24777 (28.1%) |
| ≥2010 | 2074 (71.9%) | 367895 (66.3%) | 5341 (74.4%) | 605461 (67.8%) | 4471 (49.9%) | 509443 (62.8%) | 1741 (70.8%) | 326886 (64.3%) | 756 (78.9%) | 98440 (72.7%) | 1402 (72.5%) | 29831 (71.6%) | 565 (77.1%) | 6701 (70.6%) | 1815 (79.5%) | 63493 (71.9%) |

**Supplemental Table 2.**
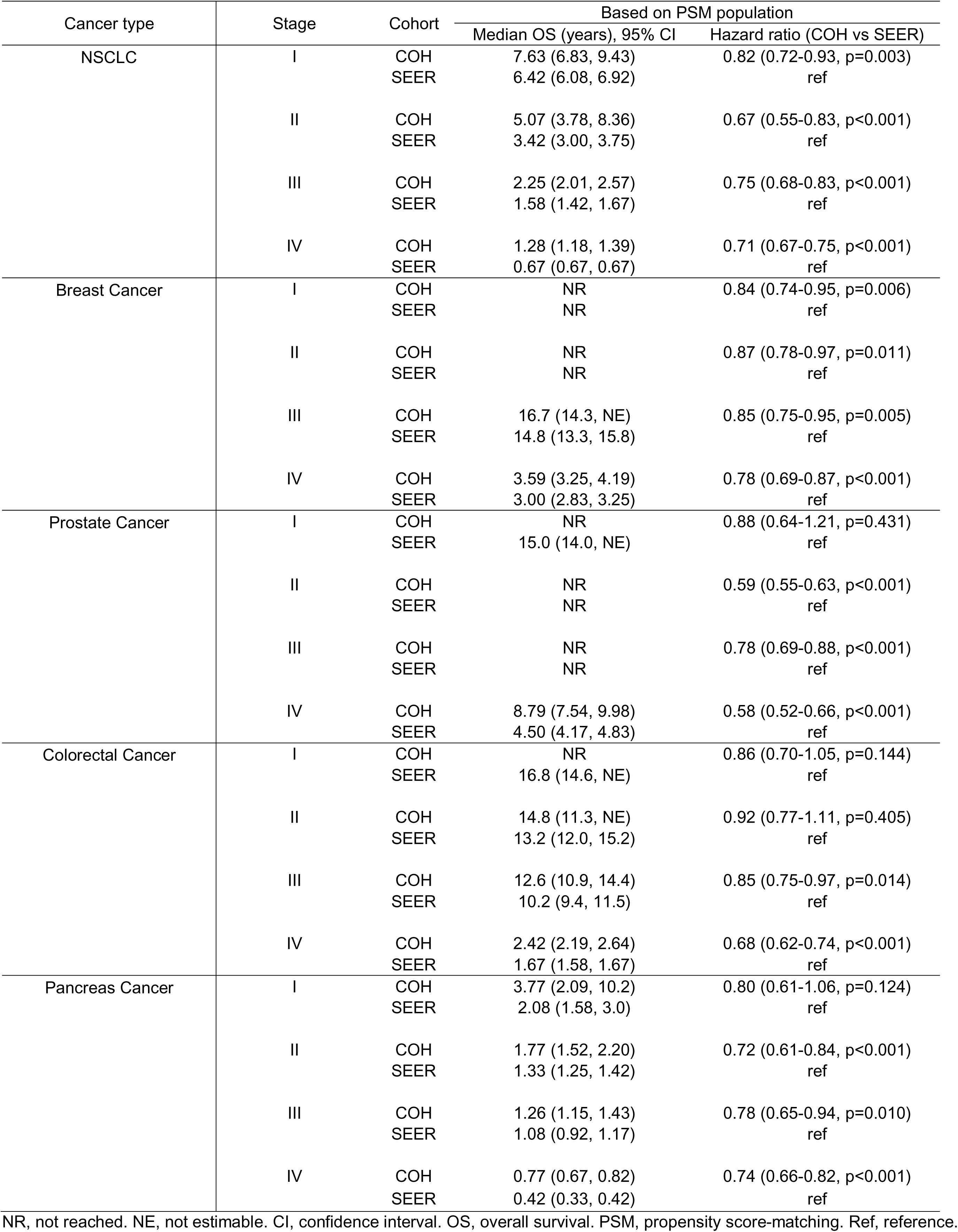
Comparison of Median Overall Survival and Hazard Ratios Between City of Hope and SEER Cohorts for Various Stages of Solid Tumor.

**Supplemental Table 3.**
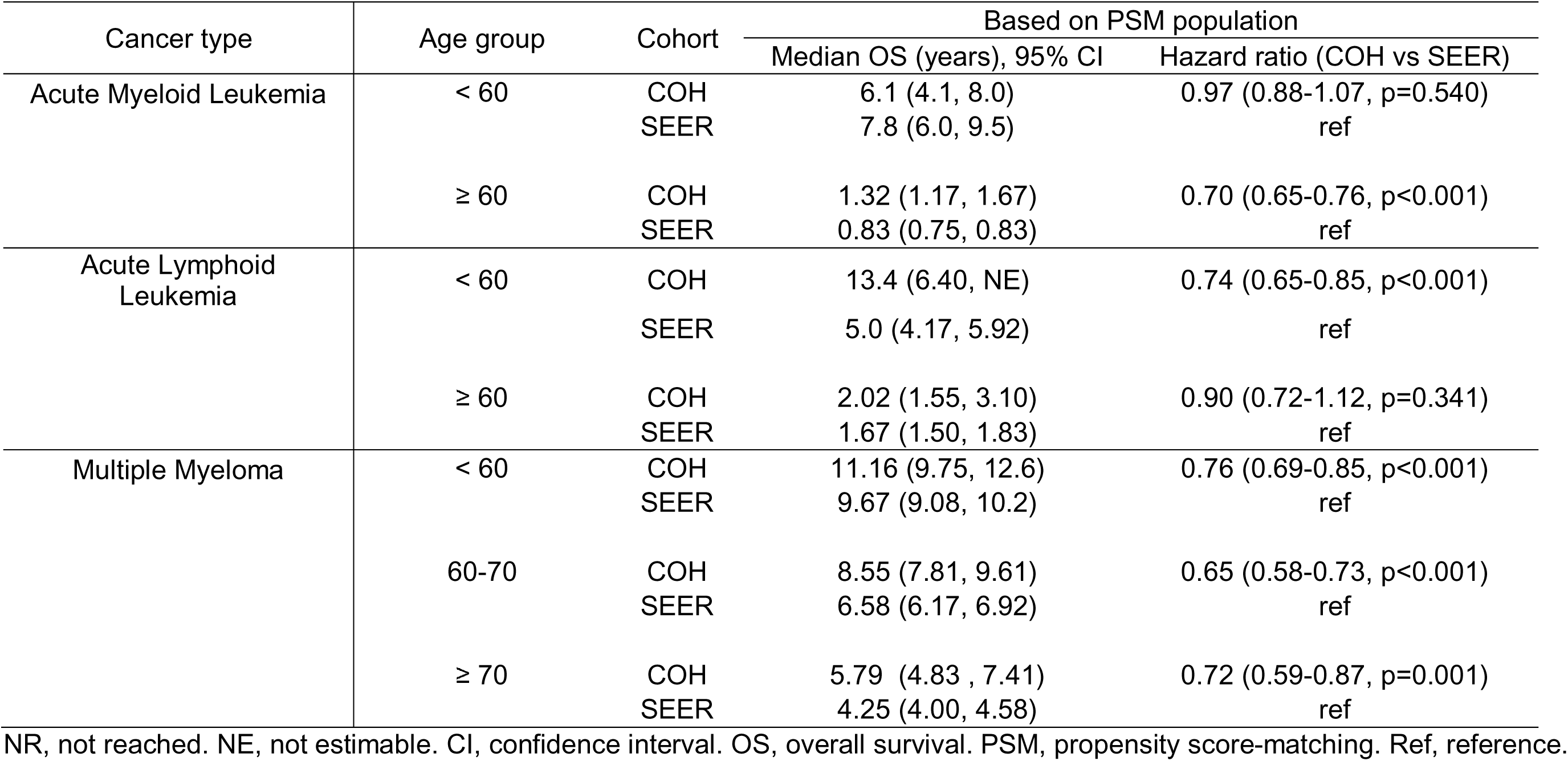
Comparison of Median Overall Survival and Hazard Ratios Between City of Hope and SEER Cohorts for Various Age Groups of Heme Malignancies.

