## Supplementary figures and images for "Cancer Survival at a Comprehensive Cancer Center Compared with Surveillance, Epidemiology, and End Results (SEER) Estimates"

### Supplemental Figure 1

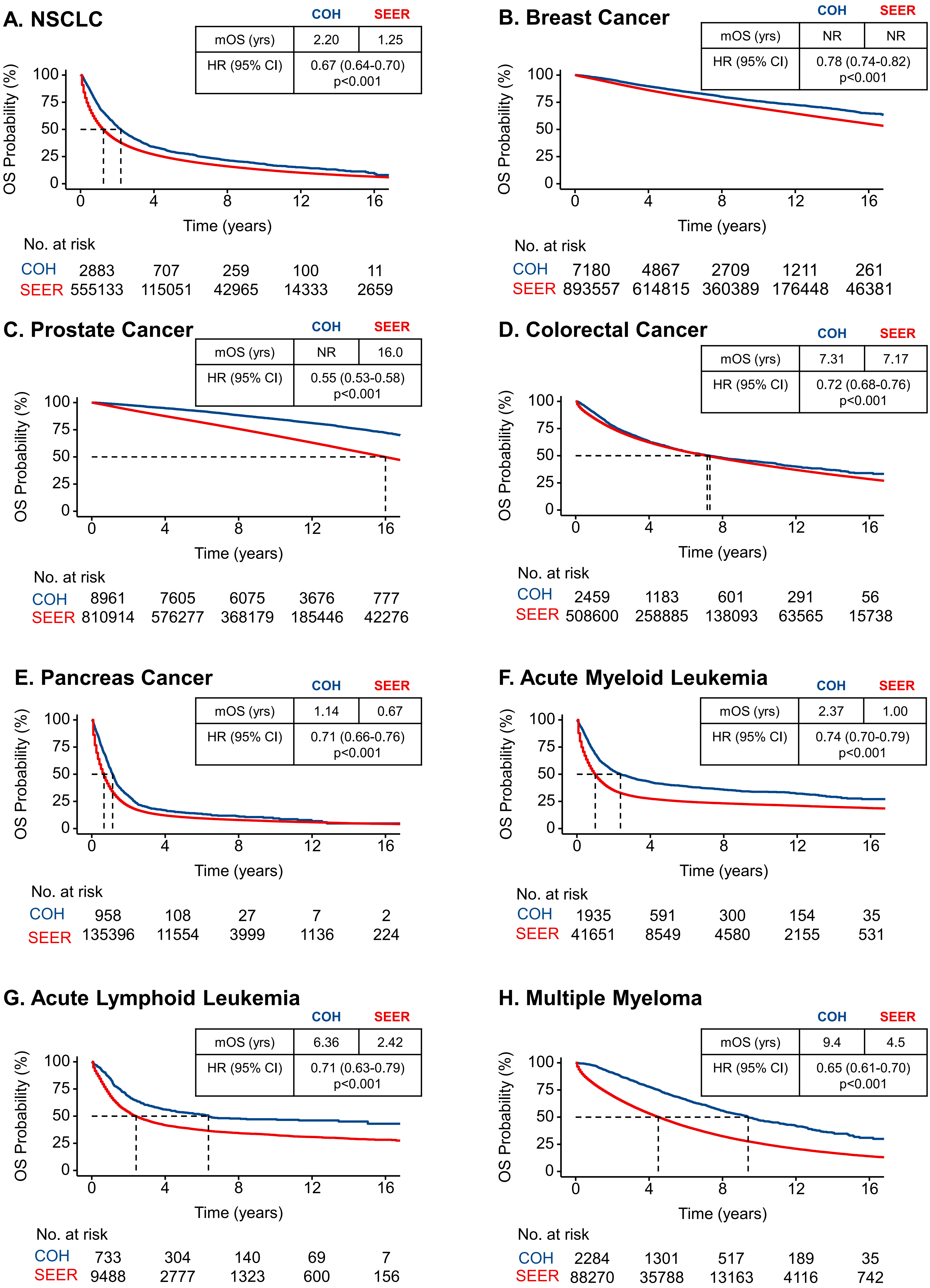

### Supplemental Figure 2

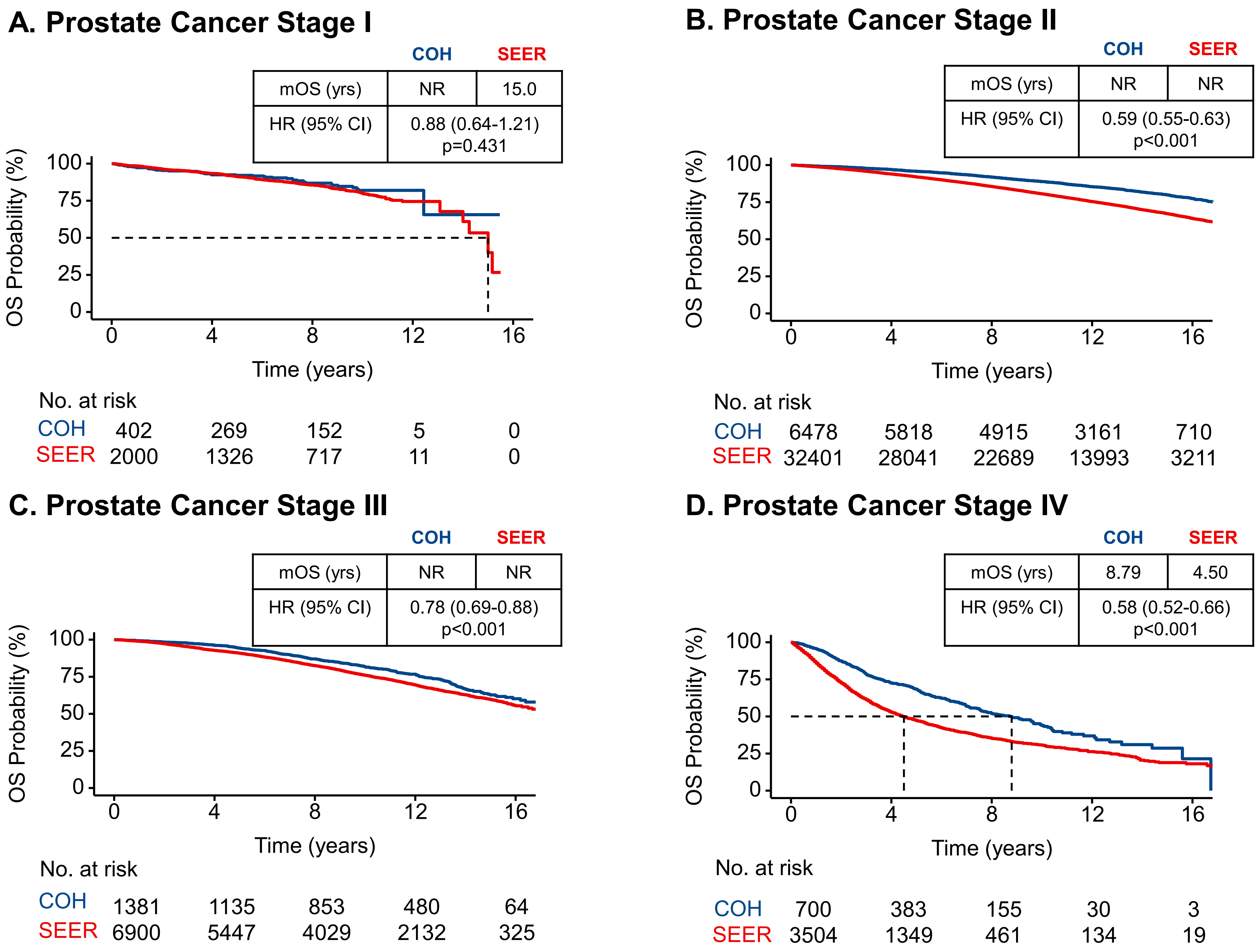

### Supplemental Figure 3

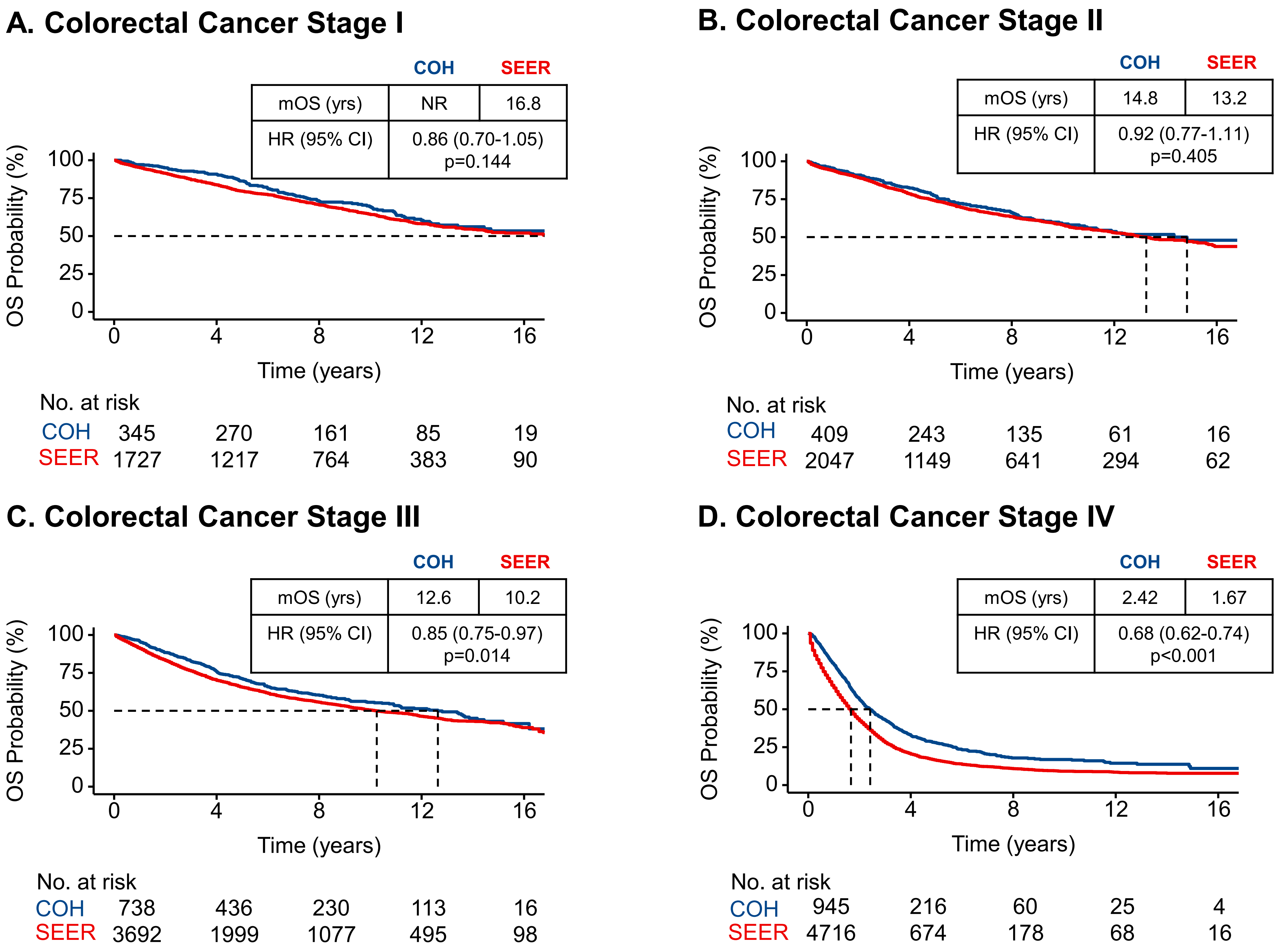

### Supplemental Figure 4

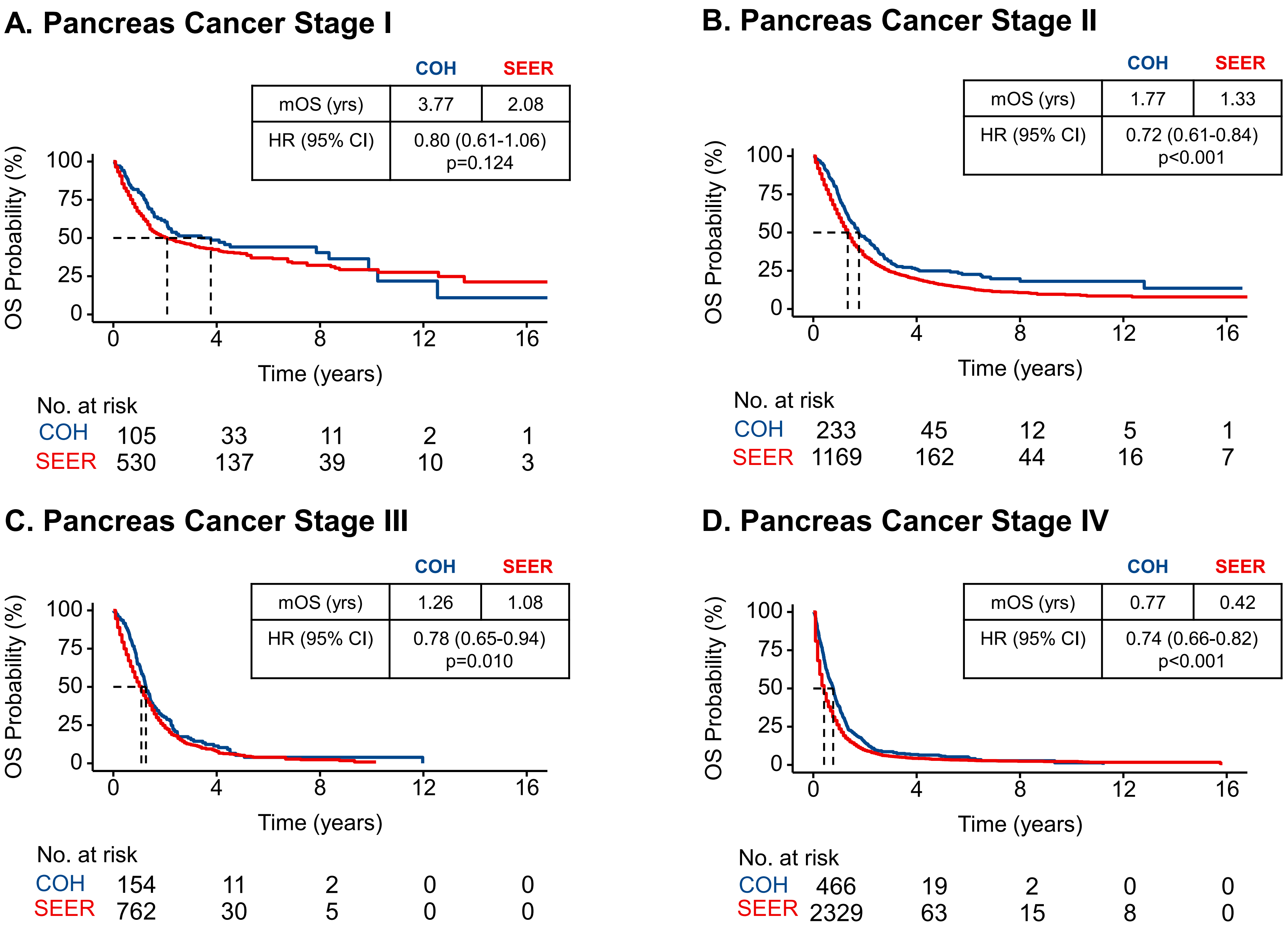

### Supplemental Figure 5

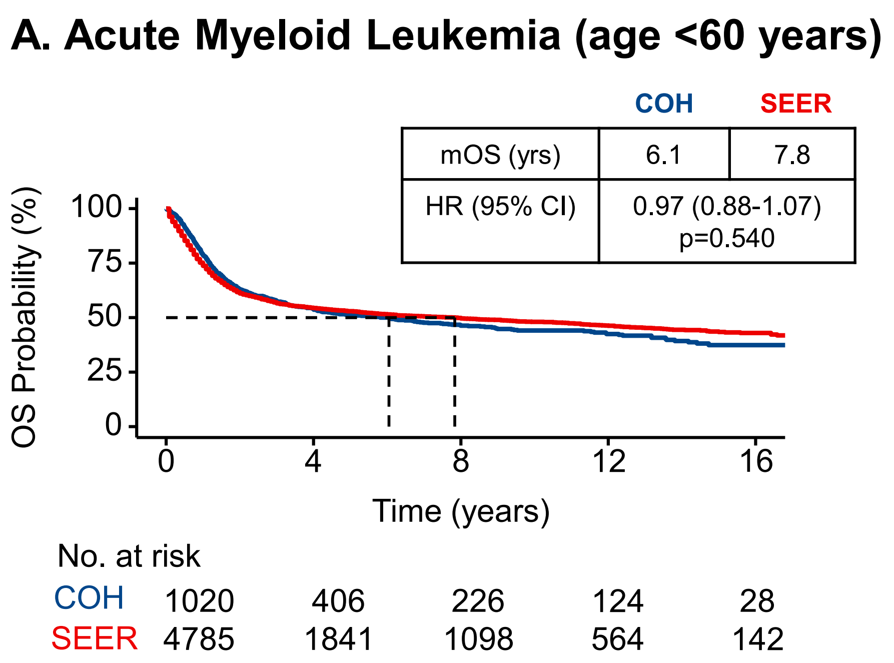

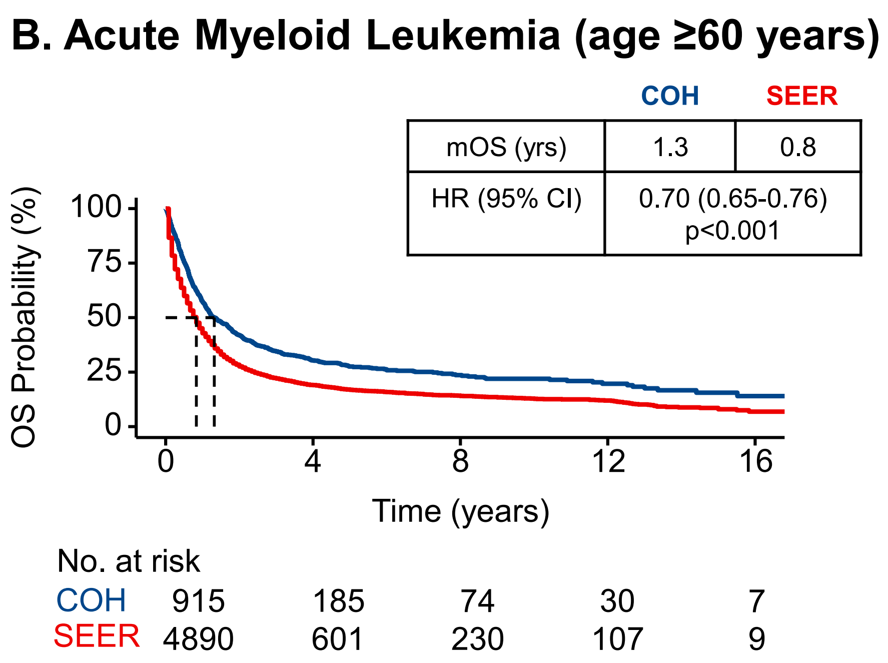

### Supplemental Figure 6

#### A. Acute Lymphoid Leukemia (age <60 years)

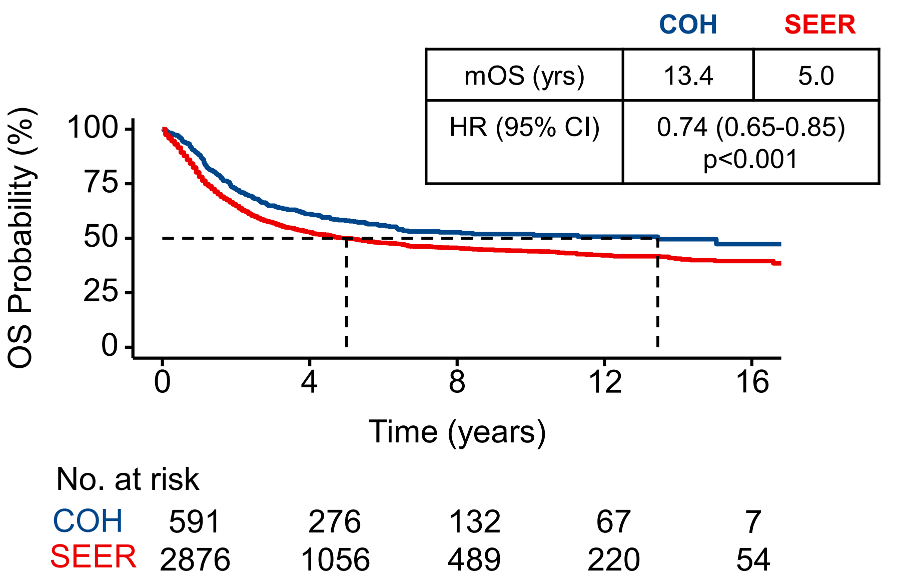


#### B. Acute Lymphoid Leukemia (age ≥60 years)

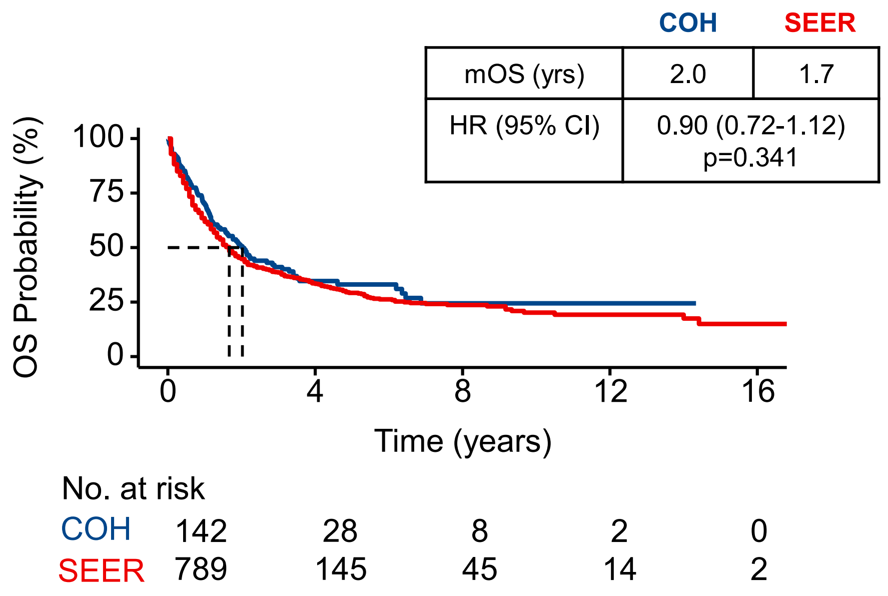

### Supplemental Figure 7

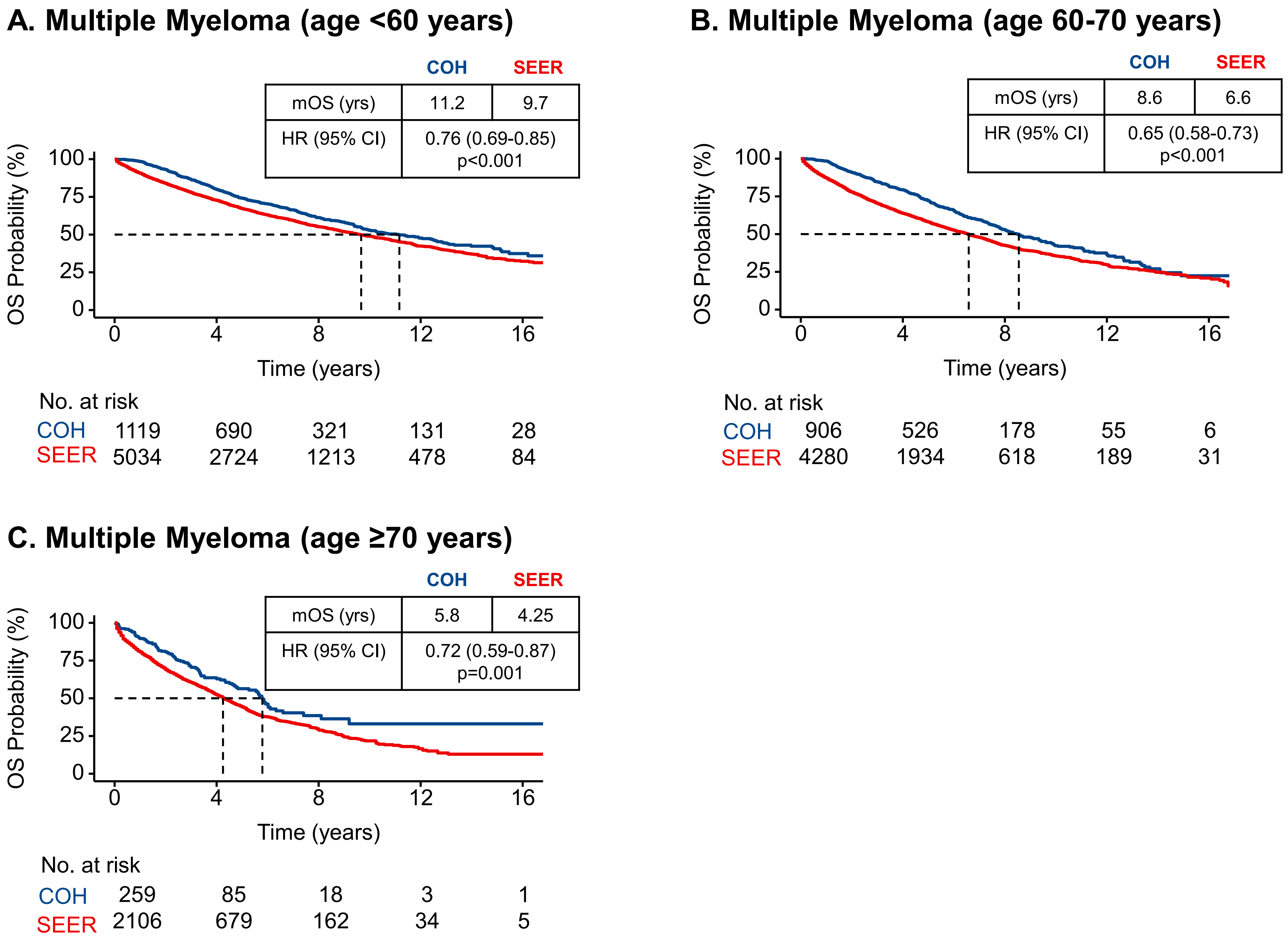
