## Supplemental Figures for "Cancer Survival at a Comprehensive Cancer Center Compared with Surveillance, Epidemiology, and End Results (SEER) Estimates"

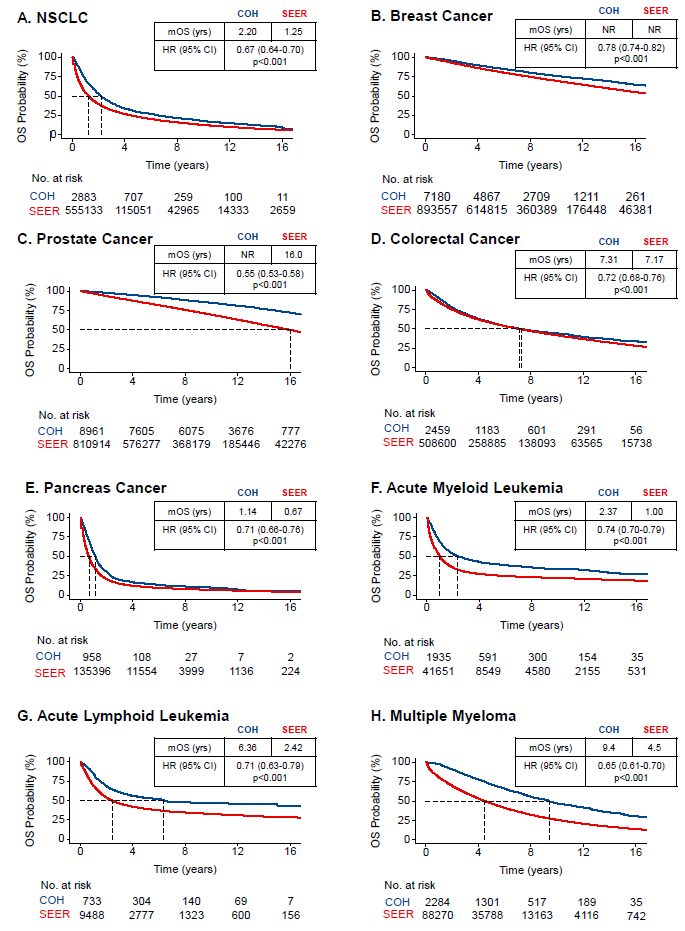

**Supplemental** **Figure 1. Overall survival comparisons after multivariable adjustment between unmatched City of Hope and SEER cohorts by Kaplan-Meier analysis for 8 cancer types.** (A) NSCLC. (B) Breast cancer. (C) Prostate cancer. (D) Colorectal cancer. (E) Pancreas cancer. (F) Acute myeloid leukemia. (G) Acute lymphoid leukemia. (H) Multiple myeloma. CI, confidence interval; HR, hazard ratio; mOS, median overall survival; NR, not reached; NSCLC, non-small cell lung cancer; OS, overall survival. Note that the mOS is from the raw data, while the HR estimates are from the adjusted model.

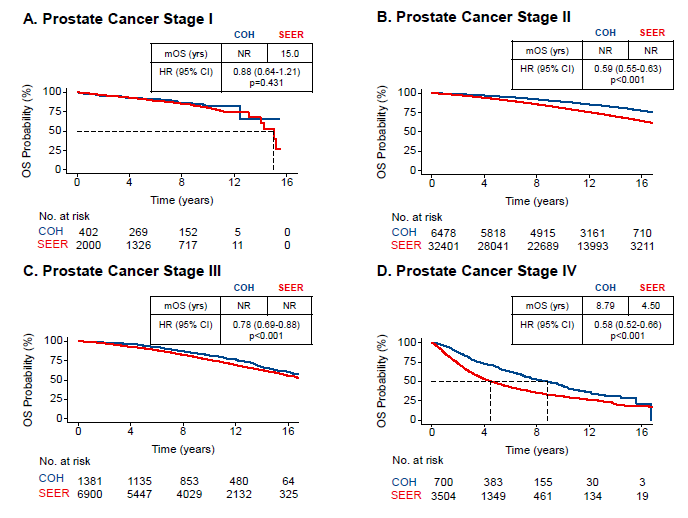

**Supplemental Figure 2. Overall survival comparisons between propensity score-matched City of Hope and SEER cohorts by Kaplan-Meier analysis for prostate cancer stages I–IV.** (A) Patients with Stage I; (B) Patients with Stage II; (C) Patients with Stage III; (D) Patients with Stage IV. CI, confidence interval; HR, hazard ratio; mOS, median overall survival; NR, not reached; OS, overall survival.

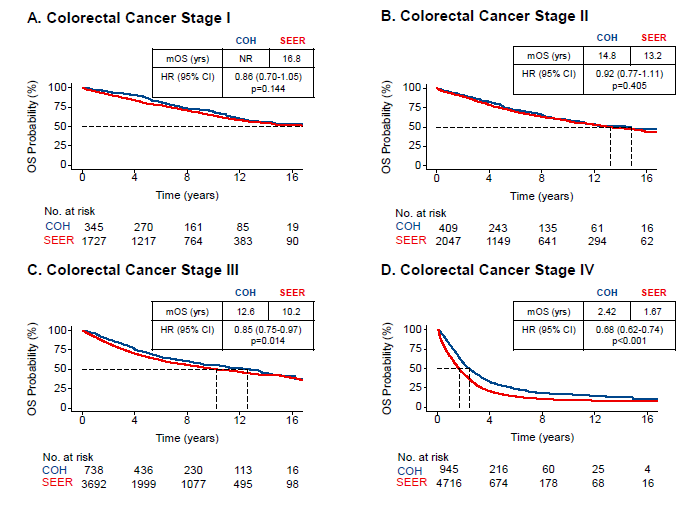

**Supplemental Figure 3. Overall survival comparisons between propensity score-matched City of Hope and SEER cohorts by Kaplan-Meier analysis for colorectal cancer stages I–IV.** (A) Patients with Stage I; (B) Patients with Stage II; (C) Patients with Stage III; (D) Patients with Stage IV. CI, confidence interval; HR, hazard ratio; mOS, median overall survival; NR, not reached; OS, overall survival.

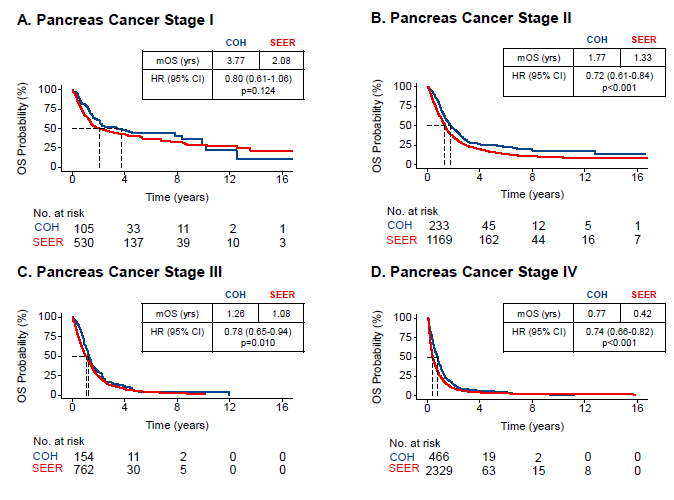

**Supplemental Figure 4. Overall survival comparisons between propensity score-matched City of Hope and SEER cohorts by Kaplan-Meier analysis for pancreas cancer stages I–IV.** (A) Stage Il; (B) Stage II; (C) Stage III; (D) Stage IV. CI, confidence interval; HR, hazard ratio; mOS, median overall survival; NR, not reached; OS, overall survival.

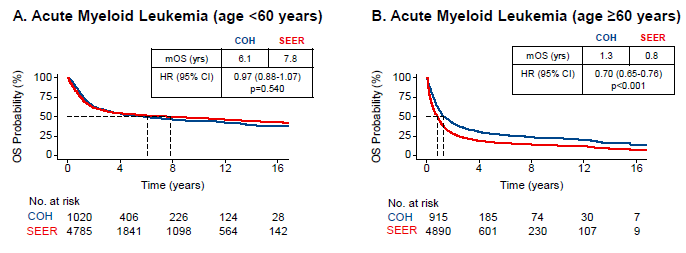

**Supplemental Figure 5. Overall survival comparisons between propensity score-matched City of Hope and SEER cohorts by Kaplan-Meier analysis for acute myeloid leukemia (AML) by age.** (A) Patients aged <60 years; (B) Patients aged ≥60 years. CI, confidence interval; HR, hazard ratio; mOS, median overall survival; NR, not reached; OS, overall survival.

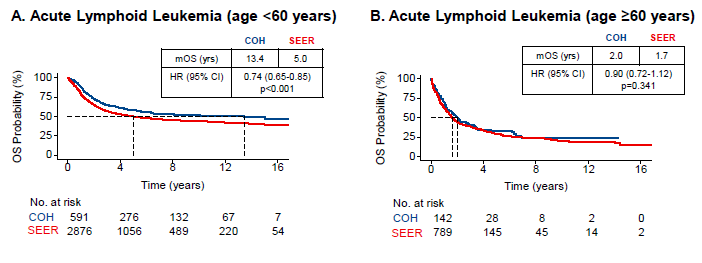

**Supplemental Figure 6. Overall survival comparisons between propensity score-matched City of Hope and SEER cohorts by Kaplan-Meier analysis for acute lymphoid leukemia (ALL) by age.** (A) Patients aged <60 years. (B) Patients aged ≥60 years. CI, confidence interval; HR, hazard ratio; mOS, median overall survival; NR, not reached; OS, overall survival.

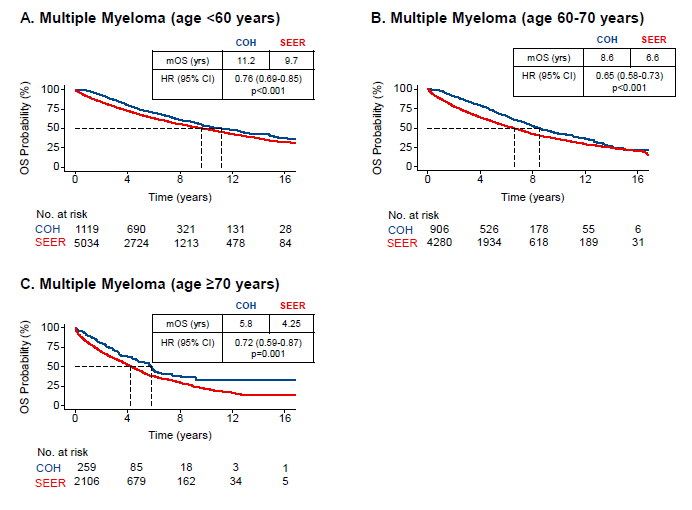

**Supplemental Figure 7. Overall survival comparisons between propensity score-matched City of Hope and SEER cohorts by Kaplan-Meier analysis for multiple myeloma by age.** (A) Patients aged <60 years. (B) Patients aged 60 to 70 years. (C) Patients aged ≥70 years. CI, confidence interval; HR, hazard ratio; mOS, median overall survival; NR, not reached; OS, overall survival.
